# Healthcare provider-targeted mobile applications to diagnose, screen, or monitor communicable diseases of public health importance in low- and middle-income countries: a systematic review

**DOI:** 10.1101/2022.11.15.22282011

**Authors:** Pascal Geldsetzer, Sergio Flores, Blanca Flores, Abu Bakarr Rogers, Andrew Y Chang

**Affiliations:** Division of Primary Care and Population Health, Department of Medicine, Stanford University; Stanford, CA; USA; Chan Zuckerberg Biohub; San Francisco, CA; USA; Center for Innovation in Global Health, Stanford University; Stanford, CA; USA; Department of Public Health and Caring Sciences, Uppsala University; Sweden; Heidelberg University Hospital, Heidelberg; Germany; Stanford University School of Medicine; Stanford, CA; USA; Department of Epidemiology and Population Health, Stanford University; Stanford, CA; USA; Stanford Cardiovascular Institute, Stanford University; Stanford, CA; USA

**Keywords:** mHealth, mobile technology, apps, digital health, communicable diseases, infectious diseases, global health, LMIC

## Abstract

Communicable diseases remain a leading cause of death and disability in low- and middle-income countries (LMICs). mHealth technologies carry considerable promise for managing these disorders within resource-poor settings, but many existing applications simply represent digital versions of existing guidelines or clinical calculators, communication facilitators, or patient self-management tools.

We thus systematically searched PubMed, Web of Science, and Cochrane Central for studies published between January 2007 and October 2019 involving technologies that were mobile phone- or tablet-based; able to screen for, diagnose, or monitor a communicable disease of importance in LMICs; and targeted health professionals as primary users. We excluded technologies that merely digitized existing paper-based tools or simply facilitated communication. Extracted data included disease category, pathogen type, diagnostic method, intervention purpose, study/target population, sample size, study methodology, development stage, accessory requirement, country of development, operating system, and cost. Given the search timeline and the extremely rapid turnover in the science concerning Coronavirus Disease 2019 and its novel variants, studies involving it were not included in the analysis.

Of 13,262 studies identified by the screen, 33 met inclusion criteria. 12% were randomized clinical trials (RCTs), with 58% of publications representing technical descriptions. 62% of studies had 100 or fewer subjects. All studied technologies involved diagnosis or screening steps; none addressed the monitoring of infections. 52% focused on priority diseases (HIV, malaria, tuberculosis), but only 12% addressed a neglected tropical disease. Although most reported studies were priced under 20USD at time of publication, two thirds of the records did not yet specify a cost for the study technology.

We conclude that there are only a small number of mHealth technologies focusing on innovative methods of screening and diagnosing communicable diseases potentially of use in LMICs. Rigorous RCTs, analyses with large sample size, and technologies assisting in the monitoring of diseases are needed.

**Author Summary:** Although significant progress has been made in decreasing their worldwide impact, infectious diseases still represent a considerable burden of disease and death. This is especially the case in certain regions of low- and middle-income countries, where limited healthcare resources, personnel, and facilities can make it difficult to provide high quality care. Mobile health (mHealth) technologies are disruptive tools that hold considerable promise in these resource-constrained settings by circumventing some of the aforementioned obstacles.

To better understand the availability and characteristics of mHealth technologies for use in low- and middle-income countries, we systematically searched for studies published in English between January 2007 through October 2019 to identify all existing mobile phone- or tablet-based innovations targeted at healthcare providers for use against infectious diseases in these settings and summarized their qualities and performance.

We found that four times as many publications focused on tools that simply made data transfer more simple than there were on new tools for detecting or monitoring diseases. Few studies were tested under the most rigorous scientific methods. Many diagnostic technologies we identified require specialized attachments or additional laboratory equipment that connect to the smartphone or tablet, which could make their use in some settings more challenging.

## Introduction

As of 2019, communicable diseases were still the main driver of disability-adjusted life years (DALYs) in children under ten years of age globally and were responsible for six out of the top ten global causes of DALYs (1). In 2017, 35% of the years of life lost worldwide were from communicable, maternal, neonatal, and nutrition-related disorders (2). Communicable diseases not only increase mortality and reduce life expectancy in LMICs, but they also cause significant disability, leading to loss of economic productivity in impacted communities (1). Furthermore, nearly a tenth of the global burden of non-communicable diseases (NCDs) that year were attributed to an infectious cause, with the burden quantified to be 130 million DALYs (3). Additionally, many LMICs continue to be afflicted by neglected tropical diseases such as dengue virus, Chagas disease, and schistosomiasis. These are not only unique to these regions but also endemic, remaining a major contributor to morbidity and mortality in those settings (4–6).

The persistence of communicable diseases in LMICs is thought to be due to a number of factors, including incomplete development of robust public health infrastructure, shortage of healthcare providers, and continuance of major health disparities (4). New technologies could help overcome these obstacles to further accelerating the reduction in the communicable disease burden in LMICs. For one, such technologies could enable task shifting from physicians to nurses and community health workers (CHWs) with the goal of alleviating the shortage of more highly trained healthcare worker cadres in low-resource settings. One venue for doing so involves equipping such personnel with mobile health (mHealth) technologies, whose simplified user interfaces, integrated workflow protocols, and lower costs would be ideal for extending the practice capabilities of their users (7). For example, incorporating mHealth apps in routine CHW activities has been shown to be beneficial in process improvement and technology development, standards and guidelines, education and training, and leadership and management (8). mHealth devices have already been demonstrated to improve the management of infectious diseases in many instances in low-resource settings, (9,10) as they can serve as rapid and cheap diagnostic tools (11,12). The wireless, portable aspects of many such technologies also increase the accessibility of healthcare services to patients by reducing travel time and expenses (8).

The current published literature contains many reports of apps that simply digitize existing algorithms or facilitate inter-provider or patient-provider communication. To the best of our knowledge, however, it does not offer a comprehensive, up-to-date systematic review of truly innovative, novel provider-facing mHealth technologies available for infectious disease care in LMIC settings. These include technologies such as simplified laboratory testing equipment with smart device interfaces and artificial intelligence-guided diagnostic tools. As such, we conducted a systematic review that aims to identify all existing novel mobile phone- or tablet-based innovations targeted at healthcare providers and summarize the performance of these technologies.

## Methods

### Search Strategy

We searched for all studies published in English from January 2007 through October 2019 in the following databases: Cochrane Central (searched on September 30th, 2019), PubMed (searched on October 7th, 2019), and Web of Science (searched on October 7th, 2019). The databases were queried using keywords and medical subject headings (MeSH) including those attributable to smartphones, tablets, diagnosis, screening, and monitoring. A full list of the search terms used for each database are shown in **S1 Table**. The database searches, examination of abstracts, and inspection of articles’ full-text versions were not conducted in duplicate. No restrictions were placed on study design, sample size, or publication type. Finally, the reference lists of all included studies, relevant review articles, and commentaries were screened for additional references. The search process is summarized in **Figure 1**. The review was registered in The International Prospective Register of Systematic Reviews (PROSPERO; Registration number: CRD42020193945) (13). Ethical approval was not sought from the Stanford institutional review board as the study did not constitute human subjects research and consisted only of meta-research (which is exempt by definition).

**Figure 1.**
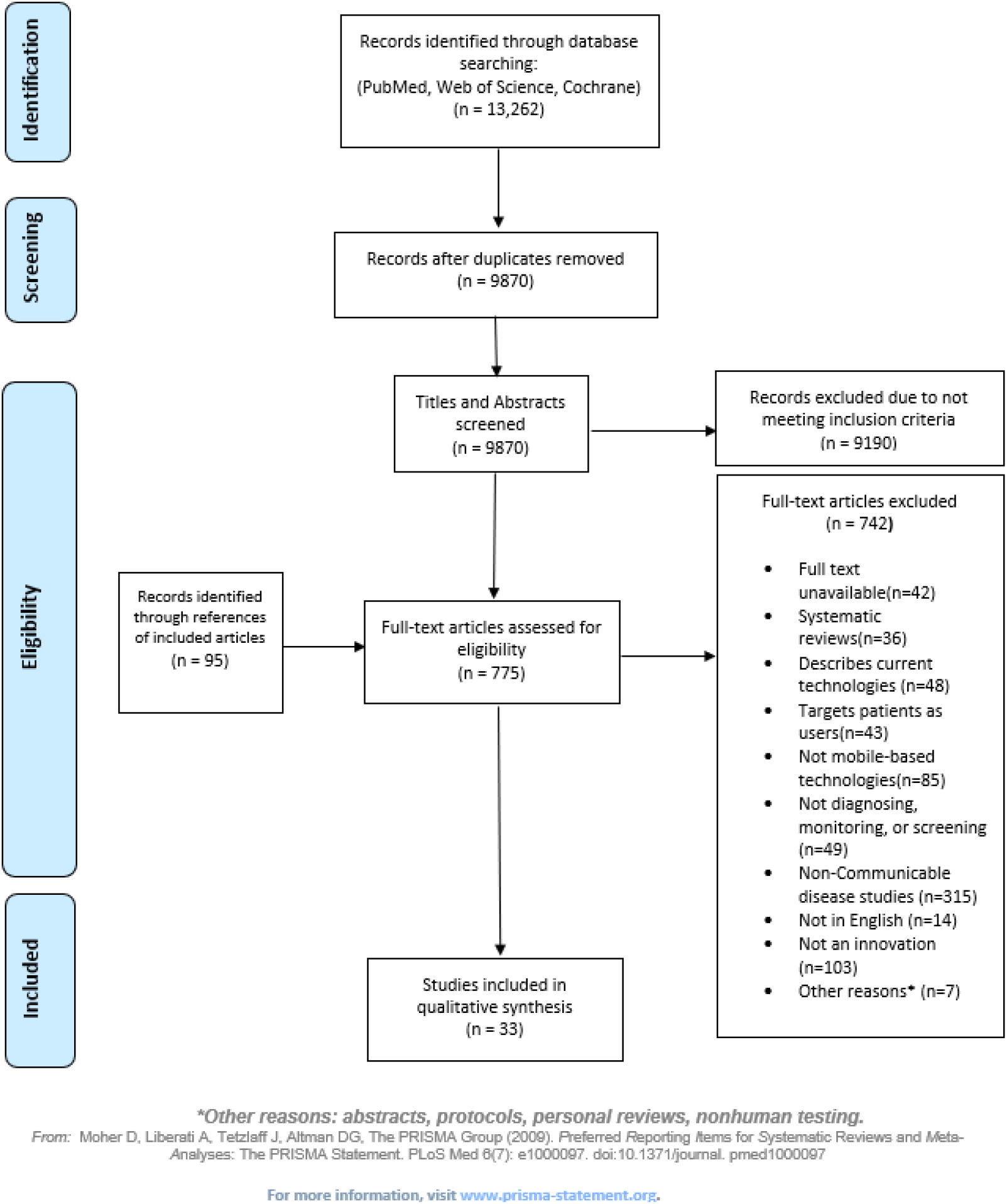
PRISMA Diagram.

## Inclusion and Exclusion Criteria

i. Titles and abstracts were screened for pertinence, and if inconclusive, the full-text versions of articles were evaluated for adequacy utilizing the following inclusion criteria:
ii. The technology reported must be mobile phone or tablet-based for their clinical function—this excludes mobile devices and applications that solely use their internet connectivity to transmit data;
iii. The technology must target healthcare professionals specifically as users— tools used to educate patients, change patient behaviors as consumer products, or improve patient-provider communication were excluded;
iv. The technology reported must be able to screen, diagnose, or monitor a disease; The technology must represent an innovation—applications simply used to keep records, reproduce existing guidelines and clinical calculators in digital form, or facilitate communication between providers were excluded; and
v. The disease the technology is designed to address must be a communicable disease of public health importance for LMICs. Such diseases were defined as infectious conditions that are estimated to cause more than 1% of deaths in any five-year age group in the general population or among neonates, or infectious diseases that have a prevalence of more than 0.1% in any five-year age group in the general population or among neonates. The Global Burden of Disease Project’s 2019 estimates were utilized for this appraisal (14). Of note, given the timeline of the search and the extremely rapid turnover in the science surrounding the disease and its many novel variants, studies involving Coronavirus Disease 2019 (COVID-19) were not included in the present analysis.

## Data Extraction

The following data were extracted from each included article: author(s), title, disease or risk factor, clinical domain by MeSH (15), intervention name, intervention type, purpose and aim of the intervention, target population, type of diagnostic method, type of pathogen studied (by microbial class, LMIC priority disease (namely Human Immunodeficiency Virus (HIV), tuberculosis and malaria), as well as neglected tropical disease (NTD) status as defined by the World Health Organization (16), type of mobile device utilized, type of software, operating system used by intervention, study population and sample size, study methods, stage of development, cost in US dollars (USD) at the time of publication (all dollar figures are given as published in the manuscript and not adjusted for inflation, and in the case of articles reporting currencies other than dollars, were converted to 2021 US dollars (17)), country of development based on first authors’ institutional affiliations, location of testing based on the study population country of residence, institutional nation of all listed authors, year of publication, and a summary of the tool (**S2 Table**). These data were extracted qualitatively using Microsoft Excel (Redmond, WA).

## Data Analysis

Quantitative data were summarized with counts and proportions. The retrieved data were organized into three themes: epidemiology, technology, and methodology. The epidemiology theme described the disease of interest (and whether it is categorized as an LMIC priority disease by the Global Health National Academies of Science (18) or diseases that were among the top ten in terms of disability-adjusted life years caused globally in 2019 (19)), its characteristics, and the geographic location of the intervention’s development. The technology theme described the primary hardware platform of the innovation, necessary peripherals, as well as the operating system it utilized and its cost considerations. The methodology theme evaluated the phase of study and research design of each publication. **S2 Table** lists these themes, as well as the categories, subcategories, and definitions that accompany them. To elucidate trends among the studies, we created tables that crossed clinical categories and included all the subthemes. We decided against conducting a meta-analysis due to the substantial degree of heterogeneity in study designs, outcome measurements, and reporting of results. As such, we employed a qualitative measure of study quality on a three-tiered (-, +, and ++) system to characterize publication quality as unsound, suboptimal, or sound, based upon the *British Medical Journal*’s Evidence Based Medicine Best Practice Toolkit (20).

## Results

Our initial search of all the above databases retrieved 13,262 results. After duplicates were removed, abstracts screened, full texts reviewed, and articles identified from reference lists of included articles were added, 33 studies met our inclusion criteria (**Figure 1, S3 Table**). Articles were excluded if they described or evaluated: i) interventions not meant for diagnosis, screening and/or monitoring (n=49); ii) non-mobile technology-based interventions (n=85); iii) interventions targeting patients instead of health professionals as users (n=43); iv) the general status of current technologies (n=48); v) non communicable diseases (n=317), or that vi) did not have a full text available (n=41); vii) were not available in English (n=14); viii) were systematic reviews (n=36); ix) were study protocols or involved non-human testing (n=8); or x) presented technology that merely digitalized protocols, scores or other procedures that could be done on paper (n=103). An overview of the included studies’ characteristics is presented in **Table 1** and the full list of identified studies is available in **S3 Table**.

**Table 1.**
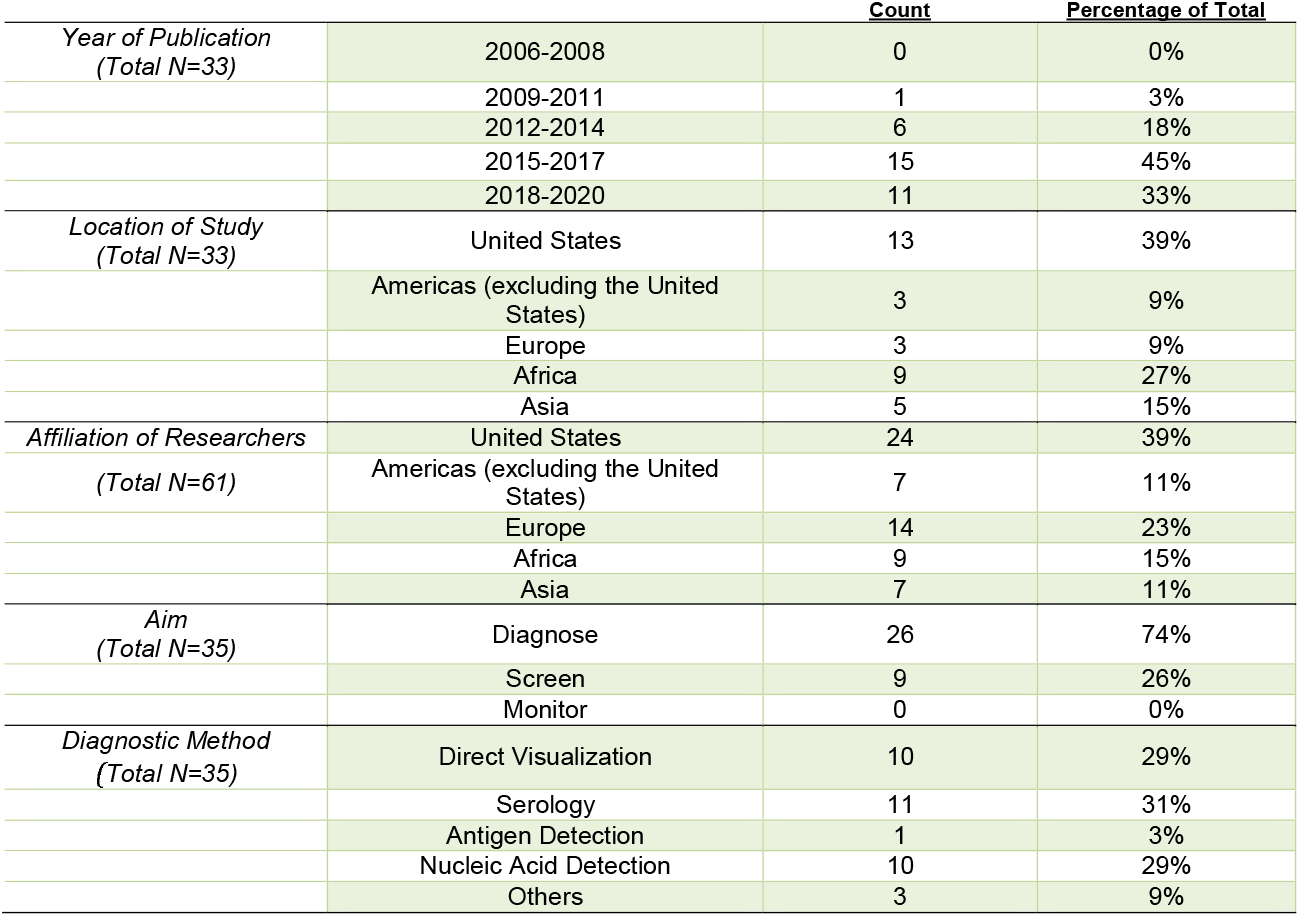
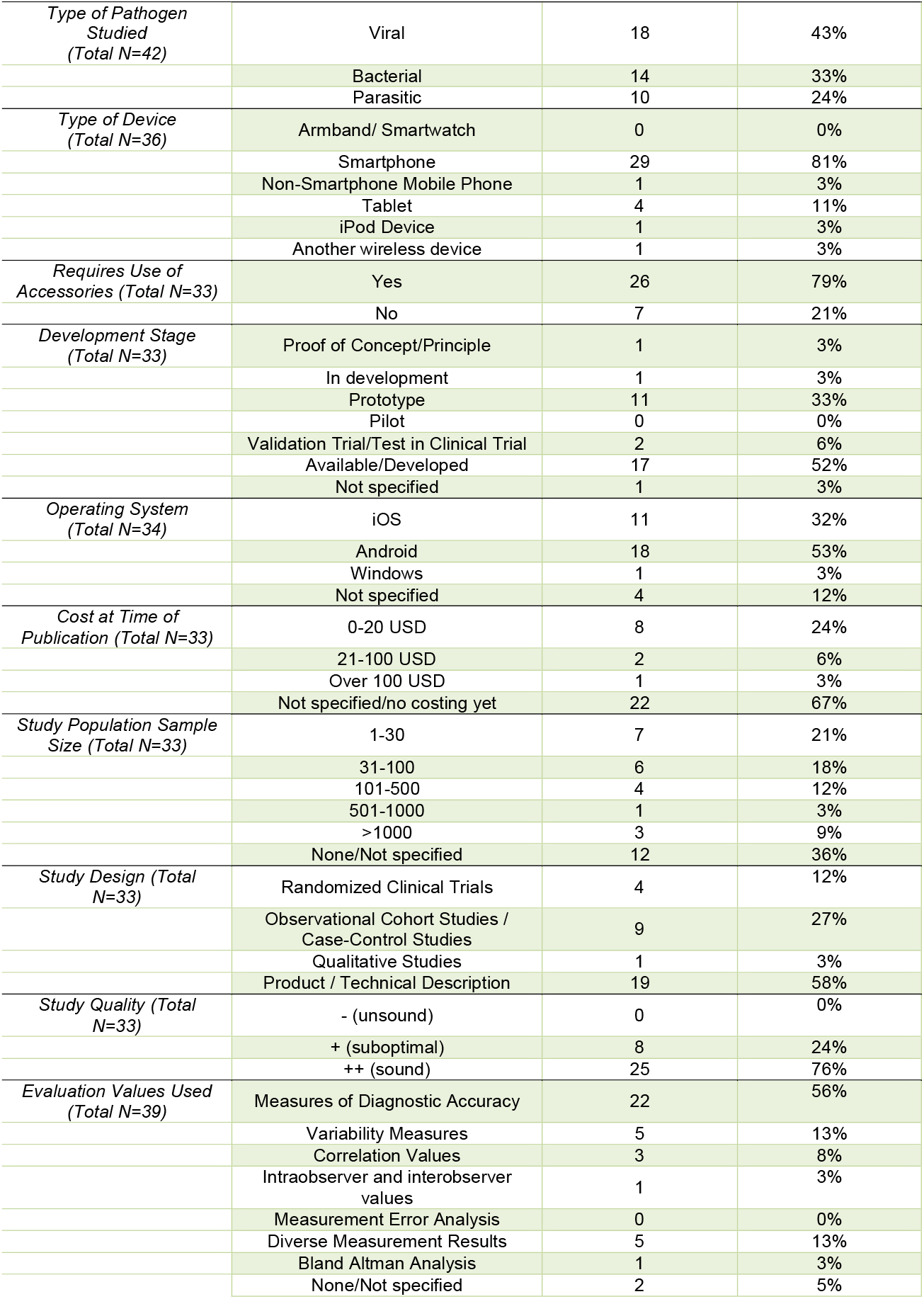
Characteristics of Studies.

## Epidemiology

Most studies described technologies tested predominantly in the United States (13/33), the rest of the Americas (3/33), followed by Africa (9/33), then Asian countries (5/33) and Europe (3/33). The affiliation of the first author’s institutions is located predominantly in the United States (24/61), with fewer based in the Americas (7/61), Europe (14/61), Africa (9/61) and Asia (7/61). A noteworthy observation is that all the studies save one (32/33) involved at least one researcher affiliated with a high-income country institution, even if the research was ultimately conducted in an LMIC.

Most of the identified technologies focus on the diagnosis of communicable diseases (26/35), while the rest aim to screen for (9/35) these diseases. No study expressed monitoring as the main aim of their technology. The diagnostic method of choice chosen by the researchers was most often serological methods (11/35), followed by direct visualization of the microorganisms (10/35) and nucleic acid detection (10/35). The technologies targeted viral (18/42), bacterial (14/42), and parasitic (10/42) infections.

Almost half (17/33) of the included studies addressed an LMIC priority disease. Only a small number of technologies (4/33) targeted a neglected tropical disease. **Table 2** describes the 16 studies of technologies aimed at diseases that were among the top ten in terms of disability-adjusted life years (DALYs) caused globally in 2019. Specifically, these studies targeted drug-susceptible tuberculosis, malaria, diarrheal diseases, and lower respiratory infections.

**Table 2:**
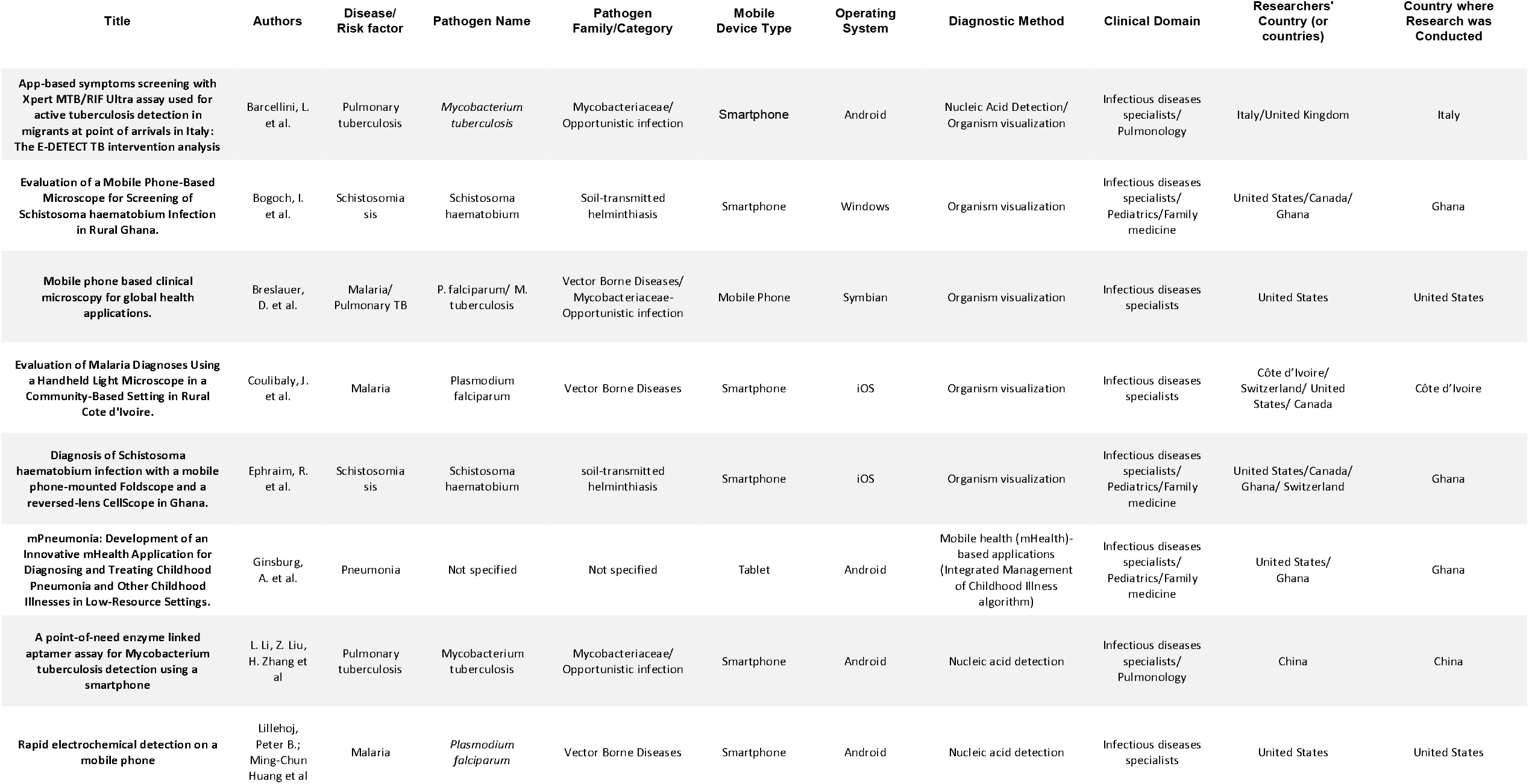
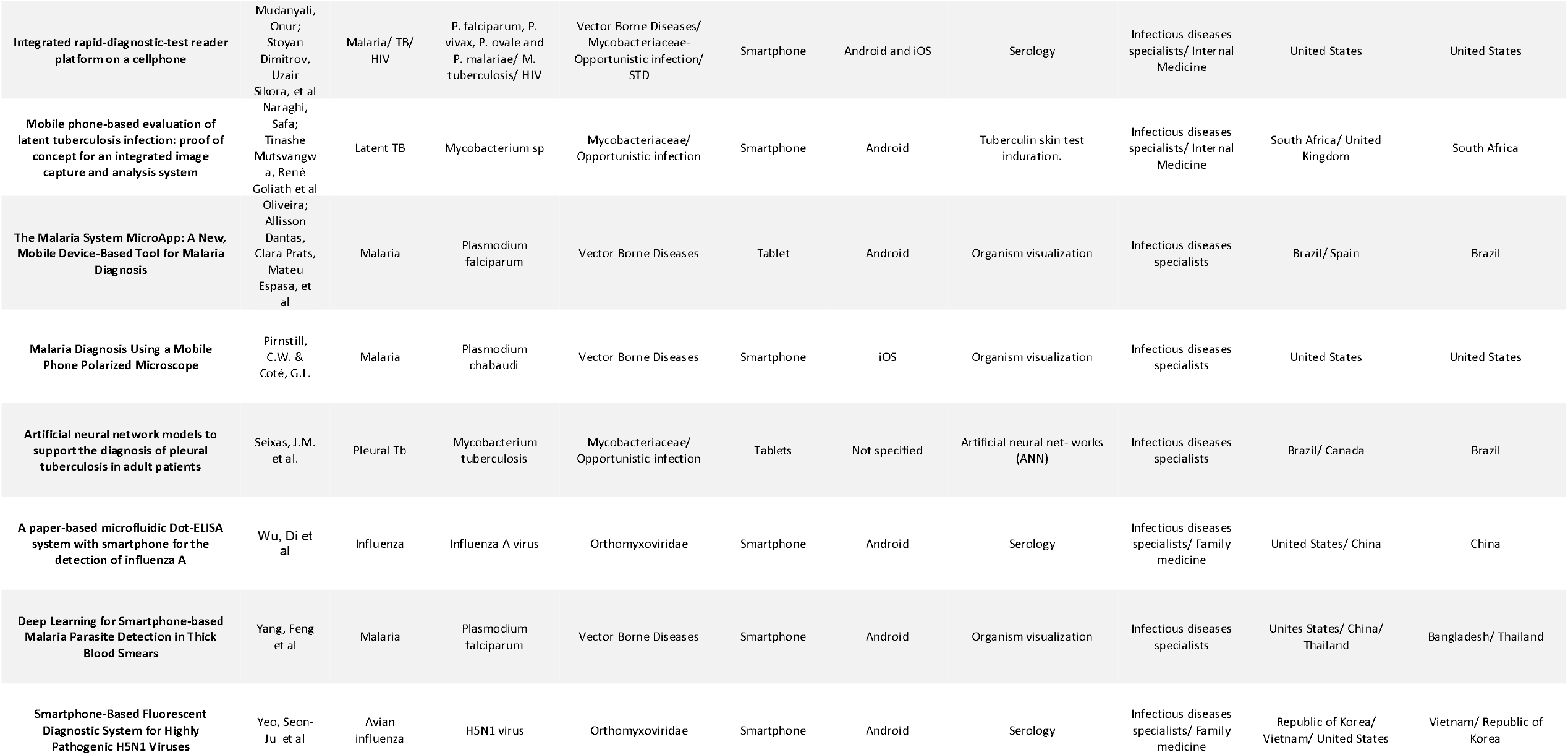
Studies of technologies addressing diseases among the top ten in disability-adjusted life years globally in 2019.

## Technology

The most popular device used in the studies was the smartphone (29/36), followed by tablets (4/36) and mobile phones without smartphone capabilities (1/36). Technologies were predominantly developed for the Android operating system (18/34) and Apple iOS (iPhone Operating System) operating system (11/34), with Windows use present in just one product (1/34). Four publications did not specify an operating system used by their application. Most (26/33) of the mobile technologies required the use of peripheral accessories attached to them such as additional optical components, 3D printed attachments, foldscopes, cradles, and dongles. Cost data were not available for most (22/33) technologies. For the technologies with costing information (11/33), most were priced at less than 20 USD (8/11), followed by between 20 and 100 USD (2/11) and one over 100 USD (1/11) at the time of study publication.

## Methodology

Most studies focused on technologies in an advanced development stage, i.e., already developed and/or commercially available (17/33) followed by studies describing prototypes (11/33). Regarding research design, most studies focused on descriptions of the technology without a formal evaluation of its efficacy or effectiveness (19/33) or assessed the technology using an observational cohort design (9/33). Only a few technologies were evaluated using a randomized design (4/33). Most publications reported study population sizes of less than 30 participants (7/33), followed by study sizes between 31 and 100 participants (6/33), then by study sizes between 101 and 500 (4/33) and over 1,000 subjects (1/33). Twelve studies did not specify a study population size.

## Discussion

### Principal Results

The aim of our study was to identify and describe mobile-based technologies targeted specifically at healthcare workers to screen, diagnose, and monitor communicable diseases of public health importance in LMICs. We focused on technologies that constituted a new tool rather than merely a digitization of an existing paper-based tool or providing a means of communicating between healthcare providers. Our screening found that there were four to five times as many publications on tools that merely facilitated communication, transferred data, or digitized an existing paper-based algorithm than there were on truly new tools for screening, diagnosing, and monitoring diseases. Additionally, we found that most technologies described in our study were tested in high-income countries using predominantly smartphones as mobile device and Android as the operating system of choice. All but one of the included studies involved at least one author affiliated with a high-income country research institution, with 42% of first authors reporting institutional affiliations in the United States or Europe.

Although half of the technologies were already at an advanced stage of development, few were tested under the rigor of large-scale randomized controlled studies. In general, the sample size was small, with 62% of the studies reporting 100 or fewer subjects. Over half of the included publications were simply technical descriptions of a product. Though most reported studies are of relatively affordable innovations (most under 20 USD), two thirds of the records did not yet specify a price point for the study technology. Most importantly, all the technologies were involved in diagnosis or screening for diseases—none were found to address monitoring of infections. We were, however, encouraged to note that half of the identified technologies focused on LMIC priority communicable diseases such as HIV, malaria and tuberculosis, although only 12% addressed a neglected tropical disease.

Controlling communicable disorders requires prompt screening, diagnosis, and monitoring of the infectious agent, both to treat the disease in the individual and to prevent its further transmission. A plethora of diagnostic tests and procedures have been available to the medical community for decades, and yet, LMICs are still burdened with high levels of communicable diseases (21). This has been partly explained by poor availability of timely, high-quality diagnostic testing. Diagnostic laboratories in LMICs are usually poorly equipped or sparsely distributed (22), limiting their ability to provide accurate and rapid information to clinicians (23). Furthermore, the costs of building and maintaining laboratories tends to be prohibitive in resource-constrained settings (22), and training specialized technical personnel requires further financial and logistic investments that are often unavailable in these countries. Our findings seem to suggest that efforts in the development of mobile technologies have also identified laboratory- and imaging-based testing as key obstacles, with approximately four out of five of our included studies focusing on diagnosis rather than screening or monitoring.

Furthermore, many of the diagnostic technologies we identified require the use of structural appendices, optical components, or specialized laboratory equipment that connect to the smartphone/tablet and its inherent software and hardware specifications. Therefore, these devices are not intended to completely replace standard diagnostic/screening tests and procedures, but rather to make them more accessible to professionals in resource-constrained settings. That said, these innovations are also limited by their disproportionate reliance on Apple iOS operating systems (as LMIC mobile devices tend to run on Google Android operating systems (24,25)) and frequent lack of large-scale rigorous evaluation in LMIC settings (24).

The rather small number of innovations in this sphere reflects the likely limited public health impact of the presently available device marketplace. Nevertheless, testing of these technologies in LMICs, the wide range of diagnostic methods employed, and the approach to a variety of emerging infectious pathogens that are being diagnosed using these devices are encouraging findings. These would seem to indicate that not only are these technologies being developed, but some are also entering a diversification phase, which may hold promise for the field (26). Future work by mHealth researchers could focus on technologies that can be scaled in a way that allows for widespread and cost-effective implementation in resource-constrained health systems, while also expanding their use to screen and monitor diseases rather than solely diagnose them.

## Limitations

Our present study has several limitations. Most importantly, we employed a restrictive set of inclusion criteria, which excluded patient-facing devices and apps which digitized communication, algorithms/guidelines, and clinical calculators. Such technologies may have important impacts on health outcomes in resource-poor settings but were outside the scope of our review. Thus, their contribution to the overall ecosystem of mHealth interventions for communicable diseases in LMICs is not available here for context. Next, the heterogeneity of the included studies regarding their results and methodological approaches precluded us from performing a meta-analysis and systematic assessment of study quality, necessitating a qualitative grading system instead. Lastly, given the timeline of the search and the rapidly changing nature of the field (and the infectious agent itself), studies on COVID-19 were not included.

## Conclusions

This systematic review found that there are only a small number of mHealth technologies that constitute novel methods of screening, diagnosing, or monitoring communicable diseases of public health importance in LMICs. Randomized trials and evaluations with large sample sizes of these technologies are still lacking, as are applications meant to monitor diseases. Additionally, most identified products require accessories or peripheral devices, and a majority rely on operating systems not common in LMICs, thus likely precluding more widespread clinical use in these settings.

## Supporting information

PRISMA Checklist for SLR/MA

## Data Availability

All included data were generated from the published literature, with individual manuscripts the property of their authors or stakeholders.

## Acknowledgements

None.

## Author Contributions

Study concept and design: PG, SF, BF; acquisition of data: PG, SF, BF, ABR; analysis and interpretation of data: PG, SF, BF, ABR, AYC; drafting of the manuscript: PG, SF, AYC; critical revision of the manuscript: PG, SF, AYC; statistical analysis: PG, SF, AYC; obtained funding: PG

