## Supplemental Materials for "Healthcare provider-targeted mobile applications to diagnose, screen, or monitor communicable diseases of public health importance in low- and middle-income countries: a systematic review"

**Table S1**

| **Date of search** | **Database** | **Dates range** | **Search strategy** |
| --- | --- | --- | --- |
| 07.10.2019 | Pubmed | 2007-2019 | 2007:2019 [dp] (smartphone [mesh] OR smartphone [tiab] OR smart phone* [tiab] OR mobile phone* [tiab] OR mobile device* [tiab] OR cell phone* [tiab] OR tablet [mesh] OR tablet [tiab] OR tablets [tiab] OR "portable device" [tiab] OR ipad OR iphone* OR "android" OR ios) AND ("app" [tiab] OR "apps" [tiab] OR application [tiab] OR applications [tiab] OR mobile application* [tiab] OR software [tiab] OR tool [tiab]) AND (diagnosis [mesh] OR diagnose [tiab] OR diagnosed [tiab] OR diagnosis [tiab] OR diagnoses [tiab] OR diagnostic [tiab] OR diagnostics [tiab] OR screening [mesh] OR screening [tiab] OR screen [tiab] OR screened [tiab] OR screens [tiab] OR monitor [mesh] OR monitor [tiab] OR monitors [tiab] OR monitoring [tiab] OR monitored [tiab]) |
| 07.10.2019 | Web of Science | 2007-2019 | (smartphone [mesh] OR smartphone OR smart phone* OR mobile phone* OR mobile device* OR cell phone* OR tablet [mesh] OR tablet OR tablets OR ipad OR iphone* OR "android" OR ios) AND TOPIC: ("app" OR "apps" OR application OR applications OR mobile application* OR software OR tool) AND TOPIC: (diagnosis [mesh] OR diagnose OR diagnosed OR diagnosis OR diagnoses OR diagnostic OR diagnostics OR screening [mesh] OR screening OR screen OR screened OR screens OR monitor [mesh] OR monitor OR monitors OR monitoring OR monitored) Timespan: 2007-2019. |
| 30.09.2019 | Cochrane Central | 2007-2019 | ((smartphone OR smart phone* OR mobile phone* OR mobile device* OR cell phone* OR tablet OR tablets OR iphone or iphones OR "android" OR ios OR ipad)):ti,ab,kw AND (("app" OR "apps" OR application OR applications OR mobile application* OR software OR tool)):ti,ab,kw AND ((diagnose OR diagnosed OR diagnosis OR diagnoses OR diagnostic OR diagnostics OR screening OR screen OR screened OR screens OR monitor OR monitors OR monitoring OR monitored)):ti,ab,kw |

**Table S2**

| **Themes** | **Subthemes** | **Categories** | **Definitions or Subcategories** |
| --- | --- | --- | --- |
| Epidemiology | Diagnostic Method | Direct Organism visualization | Diagnostic procedure in which pathogens are visualized to evaluate morphological details and establish identity. |
|  |  | Serologic Tests | Diagnostic procedures involving immunoglobulin reactions. |
|  |  | Antigen Detection | Diagnostic procedures detecting the presence of microorganism’s antigens. |
|  |  | Nucleic Acid Detection | Diagnostic procedure assaying bodily fluids or tissues for the presence of viral or bacterial DNA or RNA. |
|  |  | Others | Tuberculin Skin Test |
|  | Microbiological Category | Virus | An infective agent that typically consists of a nucleic acid molecule in a protein coat, is too small to be seen by light microscopy, and can multiply only within the living cells of a host. |
|  |  | Bacteria | A member of a large group of unicellular microorganisms which have cell walls but lack organelles and an organized nucleus, including some that can cause disease. |
|  |  | Parasite | An organism that lives in or on an organism of another species (its host) and benefits by deriving nutrients at the other's expense. There are three main classes of parasites that can cause disease in humans: protozoa, helminths, and ectoparasites. |
|  | LMIC Priority Diseases | Yes/No | HIV/AIDS, tuberculosis, and malaria are considered priority communicable diseases in LMICs and are often analyzed as such given their funding status with the Global Fund |
|  | Neglected Tropical Disease (NTD) | Yes/No | The WHO/CDC recognizes the following diseases as NTDs: Chagas disease, Buruli ulcer, Dengue & Chikungunya, Dracunculiasis, Echinococcosis, Fascioliasis, African trypanosomiasis, Leishmaniasis, Leprosy, Lymphatic filariasis, Onchocerciasis, Rabies, Schistosomiasis, Soil-transmitted helminthiasis, Cysticercosis, Trachoma, Scabies and other ectoparasites, Mycetoma/deep mycoses, Yaws, and Snakebite envenoming |
| Technology | Type of Mobile Device* | Armband/Smartwatch | Wearable computers in the forms of a watch or a band usually wrist worn. |
|  |  | Smartphones | A cell phone that includes additional software functions (such as email or an Internet browser) |
|  |  | Mobile Phones | A portable, usually cordless, telephone for use in a cellular system |
|  |  | Tablets | A mobile computing device that has a flat, rectangular form like that of a magazine or pad of paper, that is usually controlled by means of a touch screen, and that is typically used for accessing the Internet, watching videos, playing games, reading electronic books, etc. |
|  |  | iPod Devices | A small electronic device for playing and storing digital audio and video files, proprietary of Apple Inc. |
|  |  | PC | A general-purpose computer equipped with a microprocessor and designed to run especially commercial software (such as a word processor or Internet browser) for an individual user |
|  |  | Other wireless devices | Blue Box device, Bluetooth enabled monitoring system, digital camera, UWB P410 Radar Module, wireless bio patch |
|  | Development Stage | Proof of Concept/Proof of Principle | Evidence that shows that a (…) design idea, technology etc. will work, usually based on an experiment or a pilot project. |
|  |  | In development | Technology in the process of being prepared, developed or completed. |
|  |  | Prototype | A first full-scale and usually functional form of a new type or design of a construction |
|  |  | Pilot | A small-scale preliminary study conducted to evaluate feasibility, duration, cost, adverse events, and improve upon the study design prior to performance of a full-scale research project or technology deployment. |
|  |  | Validation Trial/Test in Clinical Trial | A scientifically controlled study of the safety and effectiveness of a therapeutic agent (such as a drug or vaccine) using consenting human subjects |
|  |  | Available/Developed | Technologies that have undergone development and trials and are currently available to be used by practitioners, including those available at virtual application stores for purchase or free download. |
|  |  | Not specified | Study does not state the stage of development. |
|  | Operating System | iOS | Mobile operating system created and developed by Apple Inc. exclusively for its hardware. |
|  |  | Android | Mobile operating system based on a modified version of the Linux kernel and other open-source software, designed primarily for touchscreen mobile devices such as smartphones and tablets |
|  |  | Windows | A group of several proprietary graphical operating system families, all of which are developed and marketed by Microsoft and targeted to different devices, ranging from Personal Computers to Mobile Phones. |
|  |  | Blackberry | A proprietary mobile operating system developed by Canadian company BlackBerry Limited for its BlackBerry line of smartphone handheld devices. |
|  |  | Others | Symbian |
|  |  | Not specified | Not detailed in the study |
|  | Use of Accessories | Yes/No | Use or not of accessories additional to the mobile technology to achieve its purpose. |
|  | Cost | Ranges of $USD | 0-20 USD, 21-100 USD, >100 USD, or Not specified or not costing assigned yet. Costs in non-USD currencies were converted to USD dollars using April 2021 rates of conversion. |
| Methodology | Country where Research was conducted/ Country where researchers are affiliated | North America (excluding the United States) | Canada |
|  |  | United States | USA |
|  |  | South America | Brazil |
|  |  | Europe | Switzerland, Turkey, United Kingdom, France, Greece, Italy, Spain |
|  |  | Africa | Cameroon, Madagascar, Nigeria, Rwanda, South Africa, Uganda, Ghana, Tanzania, Côte d’Ivoire |
|  |  | Asia | China, Thailand, Vietnam, Israel, South Korea |
|  | Year of Publication | Year Ranges | 2006-2008, 2009-2011, 2012- 2014, 2015-2017, 2018- 2020 |
|  | Study Design | Cross Sectional | An observational study that analyzed data from a population, or a representative subset, at a specific point in time |
|  |  | Experimental | Studies where researchers introduced an intervention and study the effects of the technology between carefully selected samples or individuals. |
|  |  | Mixed Methods | Studies that used both qualitative and quantitative methods. |
|  |  | Pre and Post Test | Studies where measurements are taken both before and after an intervention to the same sample. |
|  |  | Prospective Cohorts | Longitudinal cohort studies that followed over time a group of similar individuals (cohorts) who differ with respect to certain factors under study, to determine how these factors affect rates of a certain outcome. |
|  |  | Qualitative | Studies that involved collecting and analyzing non-numerical data (e.g., text, video, or audio) to understand concepts, opinions, or experiences. |
|  |  | Technical Description/Testing | Studies that described the underlying technical mechanisms about how a technology works without performing a systematic study or test to it. |
|  |  | Retrospective Observational | Studies that looked backward and examine exposures to the use of a mobile technology as a protection factor in relation to an outcome that is established at the start of the study. |
|  | Evaluation Values | Measures of Diagnostic Accuracy | Measures to evaluate the ability of a technology to detect a condition when it is present and detect the absence of a condition when it is absent: Area Under the Curve, Sensibility, Specificity, Positive and Negative Predictive Values, Diagnostic Odds Ratio, Accuracy, Half Error Rate |
|  |  | Variability Measures | Measures the dispersion of data in a dataset when evaluating or comparing performance of technologies over other standard methods. These include Mean Standard Deviation, Mean Deviation, Mean Difference, Mean Percent Error, Mean Absolute Error |
|  |  | Correlation Values | Statistical relationship between bivariate data or two random variables. These include Spearman's Rank Coefficient, Pearson's Correlation, Mean Correlation Coefficient, Lin's Concordance Coefficient |
|  |  | Intraobserver and Interobserver Values | Assessment of observer variability (repeatability and reproducibility) when using a mobile technology. These include Cohen’s Kappa coefficient, intraclass correlation coefficient, interrater reliability, Intraobserver variability |
|  |  | Measurement Error Analysis | Studies evaluating the uncertainty associated with a measurement result obtained with the mobile technologies. |
|  |  | Diverse Measurement Results | Diverse values included in results, expressed in units such as Hertz, Lumens, percentages, pH, and others that do not fit in the other categories. |
|  |  | Bland Altman Analysis | This should be also measurement error analysis |

**TABLE S3: STUDIES INCLUDED IN THE ANALYSIS**
